# Glucose, insulin, and brain health in the UK Biobank: A Mendelian randomisation study

**DOI:** 10.1101/2025.02.10.25321985

**Authors:** Victoria Garfield, Nasri Fatih, Christopher T. Rentsch, Liam Smeeth, Krishnan Bhaskaran, Nish Chaturvedi

## Abstract

Previous research suggests that HbA_1c_ and diabetes are unlikely to be causally related to brain health and dementia outcomes. With the availability of better genetic instruments for additional glycaemic-related markers (i.e., insulin resistance (IR), fasting glucose (FPG), fasting insulin (FI) and 2hr post-glucose (2hPG), there is scope to more thoroughly examine how diabetes-related mechanisms are causally related to specific neuroimaging brain health outcomes, and/or all-cause dementia and/or Alzheimer’s dementia (AD). Data were from the UK Biobank (max *n* = 349,288) with our sample consisting of an average age of 56 years (54% women). We constructed genetic instruments for 2hPG (15 variants), FPG (109 variants), FI (48 variants) and examined their relationships with neuroimaging outcomes such as total brain volume (TBV), hippocampal volume (HV), white matter hyperintensity volume (WMHV), Alzheimer’s dementia (AD) and all-cause dementia. We ran a two-sample MR and used conventional inverse-variance–weighted (IVW), weighted median estimator (WME), as well as MR-Egger intercept for horizontal pleiotropy. We found an association between 2hPG and higher risk of AD in both the IWV (OR: 1.49 [1.17, 1.90]) and WME (OR: 1.54 [1.12, 2.10]) models. However, we did not find that this association replicated in the International Genomics of Alzheimer’s Project (IGAP) data. We also observed an association between higher FI and higher TBV (β 15.25 (3.43, 27.07) in the IVW model. MR analyses of FPG and 2hPG with volumetric brain measures showed no evidence of a causal relationship with TBV, WMHV or hippocampal volume. These findings provide interesting results in how diabetes-related markers are causally related to brain health albeit the precise mechanisms through which postprandial hyperglycaemia increases AD risk and higher insulin preserves brain volumes warrant further research.

## Introduction

Epidemiological studies have long suggested that hyperglycaemia, diagnosed type 2 diabetes (T2DM) and insulin resistance (IR) strongly relate to worse brain health, importantly increasing the risk of cognitive decline and dementias (Biessels & Despa, 2018; McCrimmon et al., 2012; Ravona-Springer et al., 2010; Xue et al., 2019)

The mechanisms behind these relationships are poorly understood (Biessels et al., 2020) and we have yet to make substantial progress in the treatment of individuals who present with both diabetes and cognitive dysfunction (Srikanth et al., 2020). The nature and direction of these relationships also remain largely elusive, with some evidence pointing towards a bidirectional association (Biessels et al., 2020), which makes prevention and intervention even more difficult as this suggests a vicious cycle in which diabetes could lead to dementia, with the dementia itself then causing additional diabetes complications (Ojo & Brooke, 2015).

Mendelian randomisation (MR) is a genetic epidemiology method which helps overcome some of the core limitations of observational studies, namely residual confounding, and reverse causation (Davey Smith & Ebrahim, 2003). To date, MR studies have largely investigated Alzheimer’s dementia (AD) as an outcome, finding no causal relationship from T2DM to AD (Garfield, Farmaki, Eastwood, et al., 2021; Hagenaars et al., 2017; Østergaard et al., 2015). Pathways to cognitive decline and dementia involve a combination of vascular and neurocognitive mechanisms that may act either independently or in concert (Kisler et al., 2017; Nelson et al., 2016). Diabetes is more strongly related to the vascular pathways but there is evidence that it also has neurotoxic consequences (Fotuhi et al., 2012). A combination of vascular and neurocognitive mechanisms which either act independently or in synchrony is likely to lead to cognitive decline and ultimately, dementia (Schneider et al., 2007; White et al., 2005). In a previous MR study, we showed that HbA_1c_ and diabetes are unlikely to be the direct causal culprits underlying poorer cognitive function, brain health and both all-cause, and AD dementia (Garfield al., 2021). However, T2DM consists of distinct sub-phenotypes characterised by different grades of insulin responsiveness (Ahlqvist et al., 2018)., and HbA_1c_ has inconsistent associations with fasting and post load glucose (Ahlqvist et al., 2018).

Now, with the availability of better genetic instruments for diabetes-related mechanisms such as IR, fasting glucose (FPG), fasting insulin (FI) and 2h post-load glucose (2hPG) (Chen et al., 2021), the present study aimed to investigate whether i) IR, FPG, FI and 2hPG are causally related to specific neuroimaging brain health outcomes, and ii) IR, FPG, FI and 2hPG are causally associated with risk of all-cause dementia and/or AD.

## Methods

### Study design

Two-sample MR (which exploits genome-wide association summary statistics derived in non-overlapping samples) was used to mitigate biased results due to the ‘winners’ curse’ (the over-estimation of genetic associations which are common in one-sample MR)(Lawlor, 2016). An important advantage of using two-sample MR is that it allows sensitivity analyses to identify unbalanced (directional) horizontal pleiotropy (described under *Statistical analyses*), which is required to satisfy MR assumptions.

### Sample

Full details of the UK Biobank (UKB) cohort are published elsewhere (Sudlow et al., 2015). Briefly, UKB consists of 500,000 males and females from the general UK population, aged 40-69 years at baseline (2006-2010). Due to the low number of participants with AD diagnoses of other ancestry, the sample was restricted to those of White British ancestry. There was a maximum of 349,288 participants of white British ancestry with both genotype and all the outcomes and covariates of interest for our study (Table 1).

**Table 1:** Sample characteristics for UKB participants considered in our study (n = 349,288)

| Sample characteristics |  |
| --- | --- |
|  | Mean (SD) / N (%) / Median (IQR) |
| Age (years) | 56.7 (8.0) |
| Sex (Male) | 188,636 (54) |
| Socioeconomic deprivation score | -2.1 (4.1) |
| Smoking |  |
| Ever | 93916 (27) |
| Physical activity (number of days) | 3.6 (2.3) |
| Hippocampal volume (mm<sup>3</sup>) | 3830.0 (433.4) |
| White matter hyperintensity volume (mm<sup>3</sup>) * | 2824 (4261) |
| Body Mass Index (kg/m<sup>2</sup>) | 27.3 (4.7) |
| Systolic blood pressure (mmHg) | 138.2 (18.7) |
| Triglycerides (mmol/mol) * | 1.5 (1.11) |
| Total cholesterol (mmol/mol) | 5.7 (1.1) |
| C-Reactive protein (mg/l) * | 1.3 (2.0) |
| Stroke | 6301 (1.8) |

\*Median (IQR)

### Genotyping and quality control (QC) in UKB

487,409 UKB participants were genotyped using one of two customised genome-wide arrays that were imputed to a combination of the UK10K, 1000 Genomes Phase 3 and the Haplotype Reference Consortium (HRC) reference panels, which resulted in 93,095,623 autosomal variants (Bycroft et al., 2018). We then applied additional variant level QC and excluded genetic variants with: Fisher’s exact test <0.3, minor allele frequency (MAF) <1% and a missing call rate of ≥5%. Individual-level QC meant that we excluded participants with excessive or minimal heterozygosity, more than 10 putative third-degree relatives as per the kinship matrix, no consent to extract DNA, mismatches between self-reported sex and genetic sex, missing QC information and non-European ancestry (based on how individuals had self-reported their ancestry and the similarity with their genetic ancestry, as per a principal component analysis of their genotype).

### Outcomes: structural brain magnetic resonance imaging (MRI) and dementia

Structural brain MRI scans were performed by UKB in a subsample of participants using standard protocols, as described in detail elsewhere (Alfaro-Almagro et al., 2018). The post-processed measures derived by UKB and used in this study included: mean hippocampal volume (cm^3^) – adjusted for head size, total brain volume (cm^3^) – adjusted for head size, volume of white matter hyperintensities (WMH, mm^3^). WMH volume was log-transformed due to positive skew. The maximum number of participants with WMH volume with neuroimaging outcomes available for this study was 32,504, after exclusion of n=114 individuals who were outliers (+3SD from the mean) and did not pass genetic QC. We report results in cm^3^ for hippocampal and total brain volume, and exponentiated betas/percentages for WMH volume.

UKB provided algorithmically defined all-cause and Alzheimer’s dementia, captured using ICD-10 codes in linked Hospital Episode Statistics (HES) data, as well as from death registry data and primary care. These algorithmically defined outcomes were provided by UKB. Coded diagnoses were compared with clinical expert adjudication of full-text medical records. Details of ICD-10 and primary care Read codes are presented in Supplementary Table 6 and Supplementary Table 7. More in-depth information on the algorithm by Wilkinson et al. can be found elsewhere (Wilkinson et al., 2019).

### Statistical analyses

Analyses were performed using a combination of the *mrrobust* package in Stata, version 17, the *MendelianRandomisation* R package, using RStudio version 1.1.456 and PLINK version 2.0.

#### Selection of genetic variants for exposures: insulin resistance, fasting glucose, fasting insulin and 2h glucose

For insulin resistance (IR) we selected the same 53 genetic variants used by Merino and colleagues (Merino et al., 2017) which were part of a 2017 MAGIC Consortium genome-wide association study (GWAS) and included 188,577 individuals (Chen et al., 2021). For the remaining glycaemic exposures (FPG/FI/2hPG), we selected genetic variants from the European summary statistics from the latest and GWAS published by the MAGIC Consortium. Details are published elsewhere (Chen et al., 2021) but briefly, this GWAS consisted of up to 281,416 individuals. We selected independent single nucleotide polymorphisms (SNPs) for inclusion in our analyses at r^2^<0.01 within a 250kb region. We used the clump function in Plink 2.0 with these parameters, which yielded 109, 48 and 15 SNPs for fasting glucose, fasting insulin and 2h glucose, respectively. We aligned alleles from the MAGIC GWAS with our UKB data prior to analysis, by multiplying any misaligned variants’ beta coefficient by -1. As we did not have directly observed data for these exposures in UKB, we were unable to estimate the variance explained (R^2^) and F-statistic in our sample. We approximated the F-statistic by calculating them in the original sample from the MAGIC GWAS, as previously recommended (Garfield & Anderson, 2024). We used the ‘t-statistic’ formula which yielded F-values of 89, 49, 107 and 54 for IR, FI, FPG and 2hPG, respectively.

#### Main MR analyses

We firstly performed linear/logistic regression to examine the associations between our genetic instruments for all three exposures, and our outcomes in PLINK 2.0, adjusted for 10 genetic principal components to account for residual population stratification. Subsequently, inverse-variance weighted (IVW) MR was implemented as our main model. The IVW calculates the effect of a given exposure (e.g. diabetes) on an outcome of interest (e.g. visual memory) by taking an average of the genetic variants’ ratio of variant-outcome (*SNPàY*) to variant-exposure (*SNPàX*) relationship estimated using the same principles as a fixed-effects meta-analysis (Burgess & Bowden, 2015). We also performed standard MR sensitivity analyses, including MR-Egger regression (which provides an intercept term to indicate the presence or absence of unbalanced horizontal pleiotropy) (Burgess & Bowden, 2015) and the weighted median estimator (WME – which may yield robust estimates when up to 50% of the genetic variants are invalid)(Bowden et al., 2016). Results are presented as exponentiated ß coefficients (multiplicative effect size) for WMHV, all-cause dementia/AD risk and unit differences in hippocampal volume (cm^3^), and total brain volume (cm^3^).

### MR assumption checks

#### MR has three strict assumptions that must be met for study results to be valid

I) The association between the genetic variants for the exposure and the exposure itself must be strong and robust (this means that these associations have usually been replicated and validated via genome-wide association studies – GWAS –). *This assumption was met because our genetic variants were all from large-scale recently published GWAS*.
II) The association between the genetic variants (for the exposure) and the outcome must only be via the exposure under study, otherwise this is known as unbalanced horizontal pleiotropy and may bias MR results. *This assumption was assessed using methods detailed later in the manuscript, including MR-Egger*.
III) The genetic variant-outcome relationship is unconfounded. One way to test this is associations between the genetic variants (for the exposure) and common covariates of the relationship under study (e.g., the exposure SNPs should not be associated with factors such as smoking). *We checked this assumption by regressing multiple covariates (body-mass index, socioeconomic deprivation, systolic blood pressure, total cholesterol, triglycerides, C-reactive protein and physical activity – for which outliers >3 standard deviations were removed –, smoking and stroke) on the IR, FPG, FI and 2hPG SNPs. We applied a Benjamini-Hochberg false discovery rate (BH-FDR) of 0.05 to account for multiple testing*.

#### MR replication analyses: Alzheimer’s dementia (AD)

Replication analyses were carried out using summary statistics from the latest GWAS of AD (Bellenguez et al., 2022), which had a maximum of 111,326 AD cases (UK Biobank specific cases: AD-by-proxy, n= 46,828 and diagnosed cases, n=2,447) and 677,663 controls. AD-by-proxy cases were defined as participants who reported a family history of dementia. We extracted log(betas) and standard errors (SEs) for 14 2hPG SNPs, 107 fasting glucose SNPs, 48 fasting insulin SNPs and 53 insulin resistance SNPs.

## Results

The overall sample consisted of 349,288 participants. The participants had a mean age of 56.7 years and were more likely to be male (54%). 27% of participants ever reported smoking, 20% belonged in the most deprived group and the overall mean body mass index was 27.3. More details on the participants characteristics are shown in Table 1.

### Main MR results

#### Associations between fasting insulin (FI), fasting glucose (FPG) and 2h post-load glucose (2hPG), insulin resistance (IR), and dementia

Using MR, we found no evidence of associations between FI/FPG/IR, and all-cause or Alzheimer’s dementia (Figure 1). There was also no association between 2hPG and all-cause dementia, but we observed a strong relationship between 2hPG and Alzheimer’s dementia (Figure 1). This association indicated that elevated 2hPG was associated with 49% increased risk of AD under the IVW model, which was consistent, albeit somewhat larger when using a weighted median estimator (54%). We also observed no issues with unbalanced horizontal pleiotropy, as all the MR-Egger intercept p-values were >0.05.

**Figure 1.**
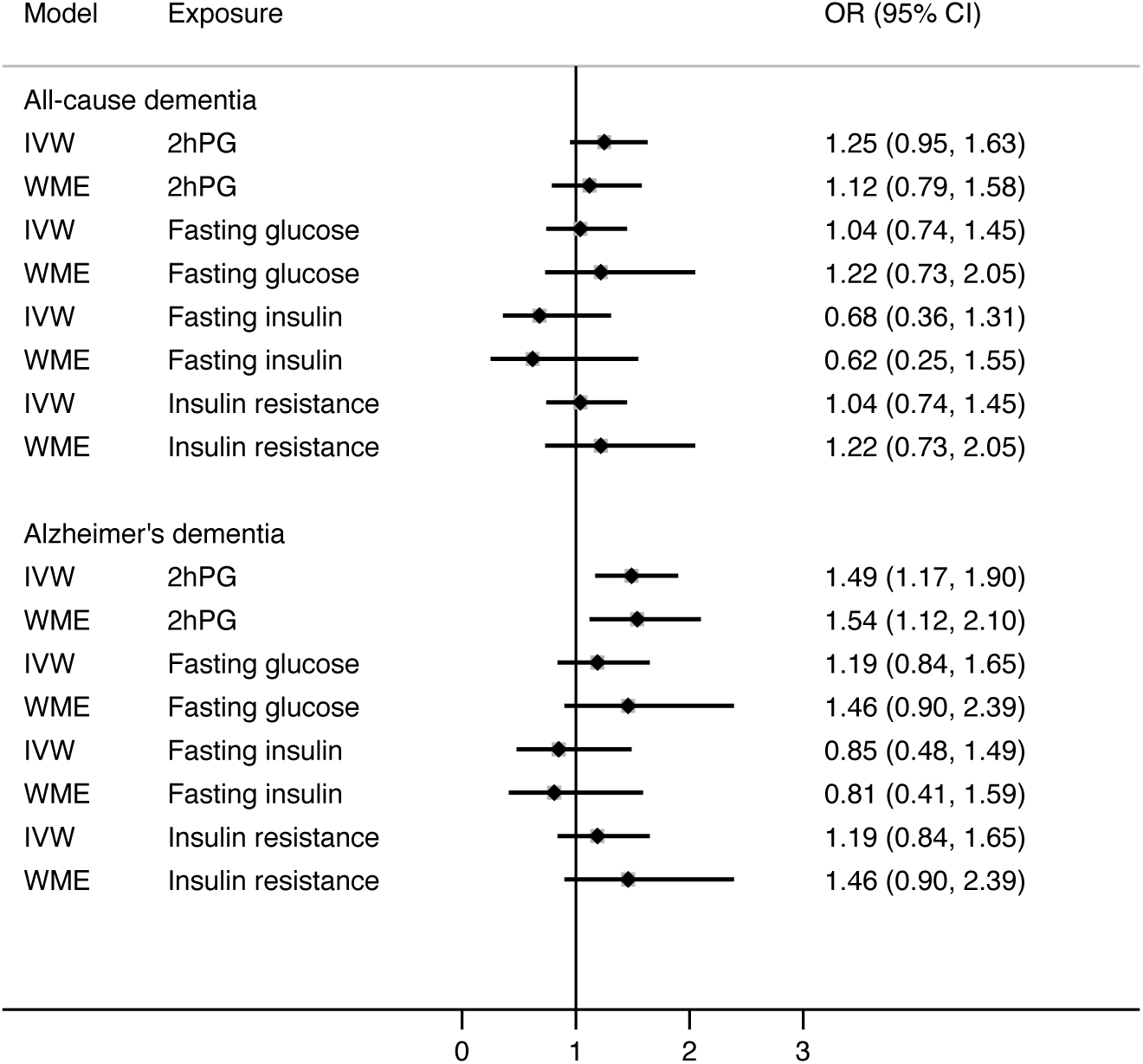
MR results for the relationship between glycaemic exposures and all-cause (N=6178), and Alzheimer’s dementia (n=2693) in UK Biobank. IVW: inverse variance weighted. WME: Weighted median estimate. 2hPG: 2-hour post-glucose. OR: Odds radio

#### Associations between FI, FPG, 2hPG, IR, and neuroimaging outcomes

We found no associations between FPG/2hPG/IR, and hippocampal volume, total brain volume or white matter hyperintensity volume (Figures 2 and 3). We did however find an association between FI and TBV, suggesting that higher insulin was associated with bigger brain volumes. All MR-Egger p-values were indicative of no unbalanced horizontal pleiotropy (>0.05).

**Figure 2.**
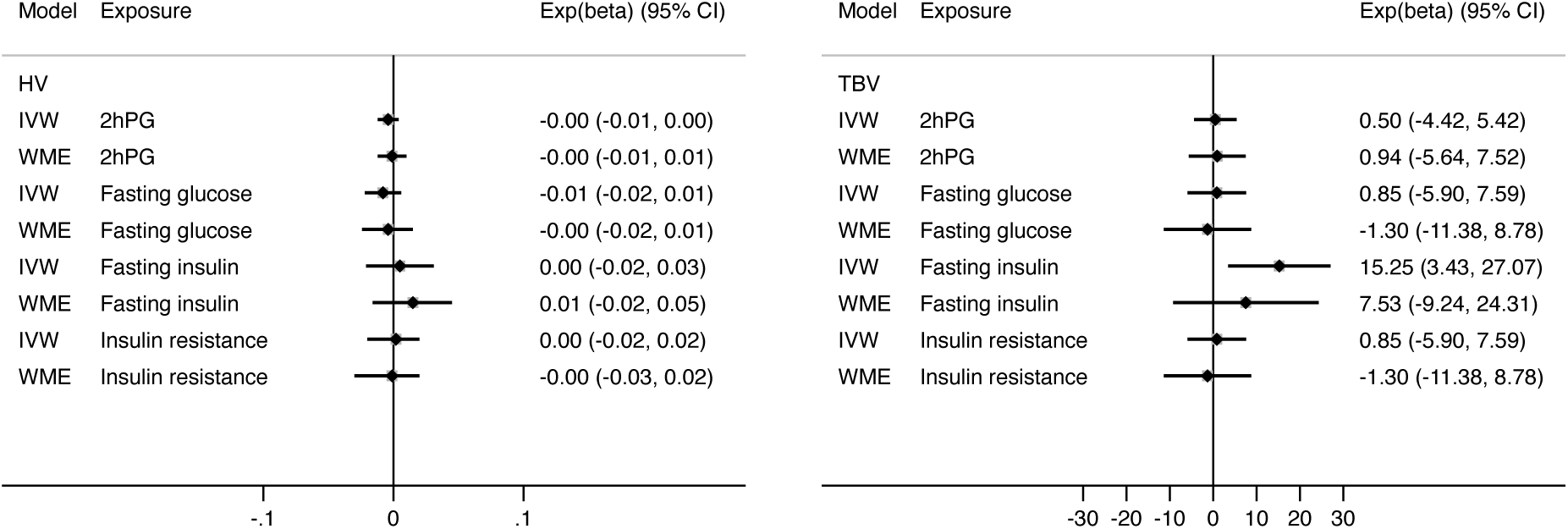
MR results for the relationship between glycaemic exposures and hippocampal, and total brain volume (n=32,504). IVW: inverse variance weighted. WME: Weighted median estimate. 2hPG: 2-hour post-glucose. HV: hippocampal volume. TBV: total brain volume.

**Figure 3.**
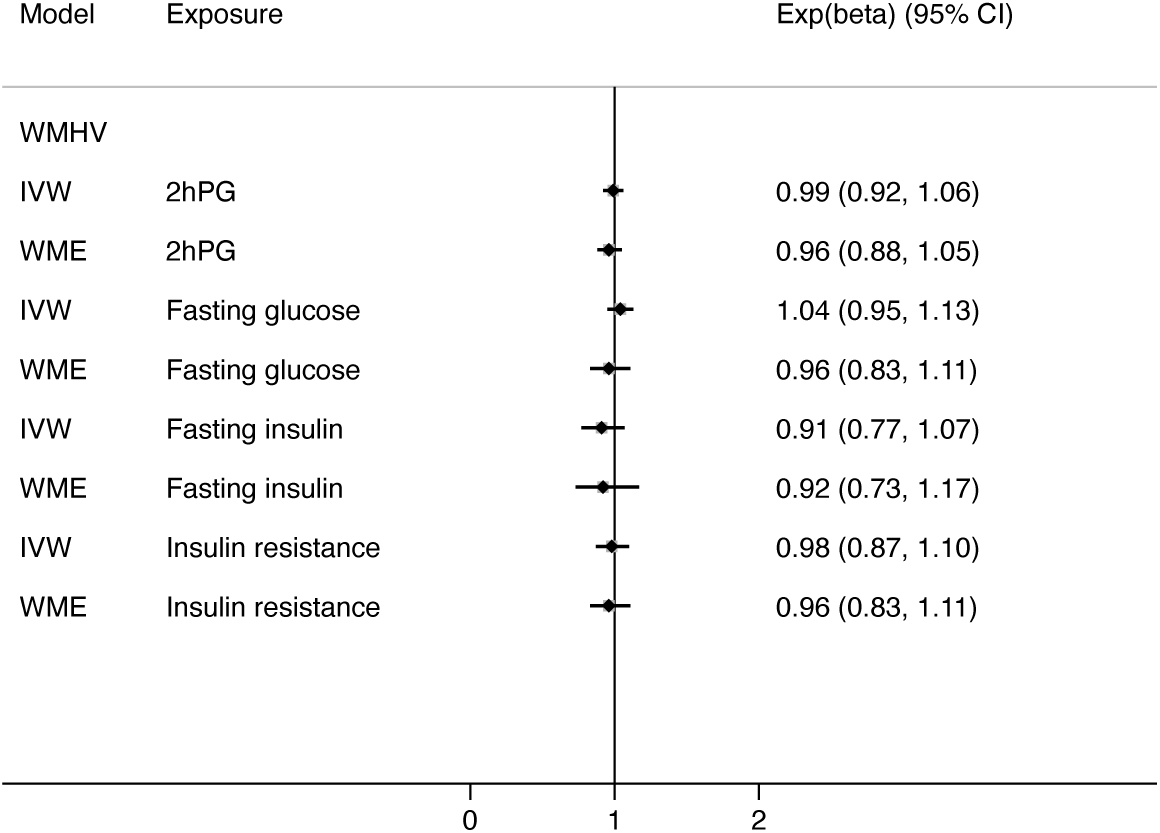
MR results for the relationship between glycaemic exposures and white matter hyperintensity volume (n=32,504). IVW: inverse variance weighted. WME: Weighted median estimate. 2hPG: 2-hour post-glucose. WMHV: white matter hyperintensity volumes.

**Figure 4.**
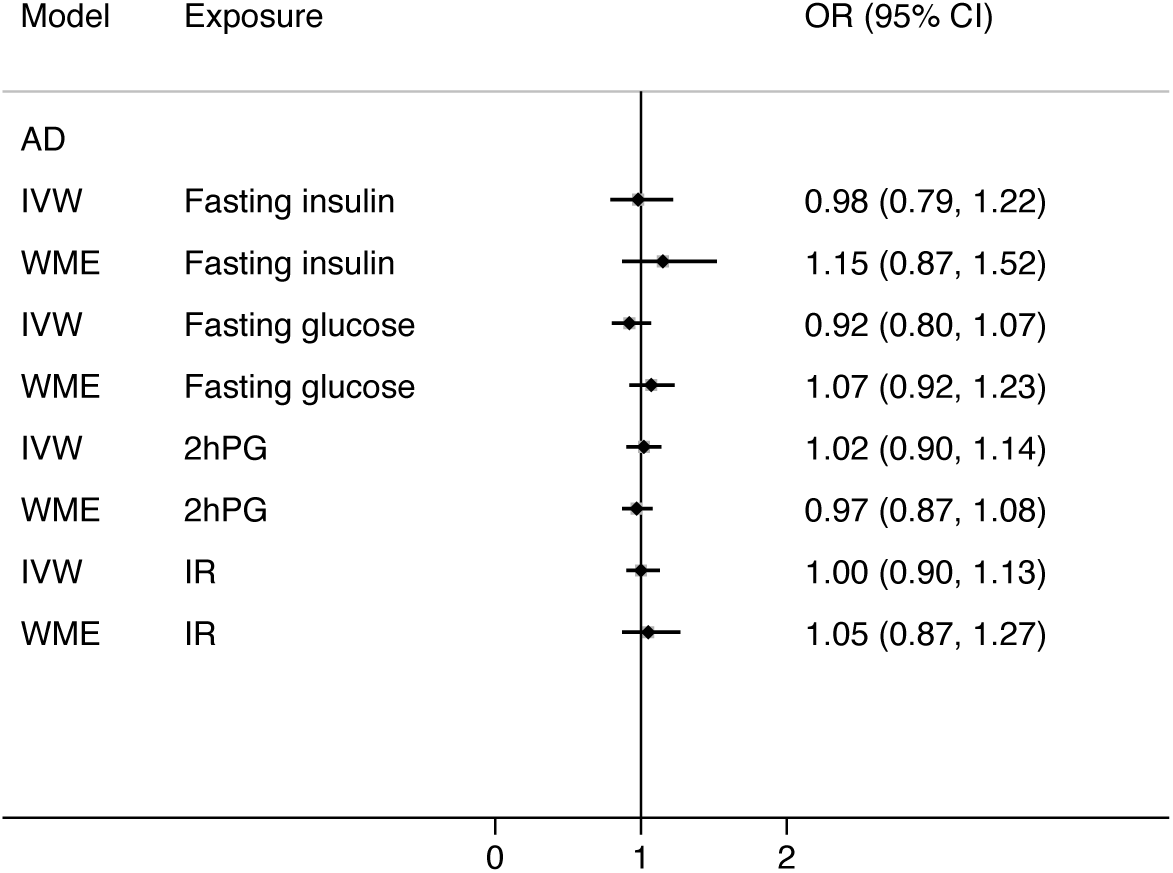
MR replication results for the association between glycaemic exposures and AD (max n_cases_=111,326). IVW: inverse variance weighted. WME: weighted median estimate. 2hPG: 2-hour post-glucose. IR: insulin resistance. AD: Alzheimer’s disease.

#### Results from MR assumption check III: associations between genetic instruments and coviates

We checked for associations between the 2hPG/FI/FPG/IR SNPs and a range of common covariates. We found associations between some 2hPG/FI/FPG/IR SNPs and CRP, triglycerides, Smoking status, SBP, body mass index, total cholesterol, physical activity and deprivation at a BH-FDR 0.05 threshold (see Supplementary tables 1-5).

## Discussion

### Overview of findings

In this two-sample Mendelian randomisation study, we found evidence of a strong association between 2-hour post-load glucose and a 49% increased risk of AD in the UKB. However, we did not find that this association replicated in a different GWAS dataset from Bellenguez and colleagues (Bellenguez et al., 2022). Our MR analyses of FPG and 2hPG with volumetric brain measures showed no evidence of a causal relationship with TBV, WMHV or hippocampal volume. In our results, however, an association between FI and larger TBV was observed.

### Findings in context

We previously published an MR study in UK Biobank and found no relationship between T2DM or HbA1c levels and measures of cognitive function and brain health (Garfield, Farmaki, Eastwood, et al., 2021). In the current analyses however, we considered additional markers of diabetes-related mechanisms and found a relationship between 2hPG and AD risk. Although this was found both using the IVW and WME approaches reducing the likelihood that the results are due to bias from invalid instruments or pleiotropy, there remains the possibility that this finding is due to chance.

Previous observational and MR studies have suggested that 2hPG is a glycaemic trait that strongly predicts poorer cardiovascular outcomes (The DECODE Study Group & on behalf of the European Diabetes Epidemiology Group, 2003; Yuan et al., 2022). Our findings suggest that the genetic predisposition for this marker of postprandial glucose is also associated with an increased risk for AD. There were some indications that the IR and FPG models also followed a similar trend to the 2hPG (i.e., higher risk of AD particularly for the WME models) albeit the confidence interval estimates of the odds ratio overlapped with 1 so this finding should be interpreted with caution.

The mechanisms through which postprandial hyperglycaemia may be related to AD are poorly understood. In this study, we failed to observed evidence of a relationship between 2hPG and both hippocampal and TBV. This perhaps suggests that the mechanism of this association may not occur through the characteristic global and/or hippocampal atrophy of AD. Future observational and MR studies should examine whether genetic tools for 2hPG are associated with the pathological markers of AD (i.e., tau and amyloid) to further shed the light on how the mechanisms of postprandial hyperglycaemia manifest in the brain. The failure to find an association between 2hPG and WMHV also suggest that this relationship may not manifest itself via vascular pathways albeit future studies should re-examine these associations with more sensitive measures of cerebrovascular disease such as those relating to microvascular pathology.

The replication analyses we carried out using summary statistics from the latest GWAS of AD failed to show a similar 2hrPG-AD association (Bellenguez et al., 2022). One explanation could be that the UKB analyses consist of a more general population sample while Bellenguez and colleagues’ cases/controls were specifically selected for dementia-related traits. Once again, such discrepancy regarding how AD was ascertained in the two different approaches could introduce heterogeneity in the data potentially diluting or altering the strength of the associations and our ability to replicate our findings in this dataset.

The failure to find a convincing association between 2hPG and FPG with the volumetric measures is largely consistent with our previous MR analyses of the relationship between HbA1c/diabetes and brain, and cognitive health (Garfield et al., 2021). Interestingly, we did find an association between higher FI and TBV although this was only observed in the IVW model. This being said, the associations between the WME and IWV were in the same directions (albeit the confidence intervals were wider in the WME). This was in contrast with IR which was not associated with TBV (or any other measures of brain health) in either model.

The observation that genetically influenced FI is associated with higher TBV, while those of IR are not, perhaps reflects the distinct biological pathways through which insulin may impact brain volumes. Previous evidence suggests that insulin modulates neuronal growth, synaptic plasticity, and neurotrophic factors especially during key developmental stages such as youth (Kleinridders et al., 2014). This perhaps raises the possibility of age dependency in these relationships with insulin being important earlier on and its effects could diminish in older adults as the brain’s developmental demands shift, and factors like IR or neurodegenerative processes become more prominent (Craft & Watson, 2004). IR reflects the body’s impaired response to insulin, a metabolic state that is typically peripheral and may not fully capture insulin’s direct effects in the brain. Such a distinction could suggest that insulin’s beneficial roles in the brain are separate from the negative consequences associated with systemic IR, highlighting a potential need to investigate the neurobiological effects of insulin beyond its metabolic functions.

Putting this result into the context of the other volumetric analyses, it could be hypothesised that fasting insulin may have a protective effect on brain health (reducing dementia risk and WMH, with higher hippocampal and total brain volumes), though the confidence intervals cross unity, making it difficult to assert this with confidence. Previous findings from the HERMES study also observed a potential protective link between insulin and heart failure although once again the robustness of the associations was questioned due to the width of the confidence intervals (Mordi et al., 2021). In middle-aged or older individuals, low FI could reflect either a failing pancreas or healthy glucose regulation, while high insulin might indicate either effective pancreas function or compensatory insulin production due to IR. These nuances suggest an important need for further analysis of insulin’s relationship with brain health rather than just dismissing it as a complete "null" result in our analyses.

Our analyses showed that MR assumptions were largely met. While we did observe associations between some of the glycaemic exposures and covariates such as CRP, triglycerides and cholesterol, such associations may be suggestive of vertical pleiotropy, where SNPs affect these factors through glycaemia, maintaining the causal chain without introducing bias. Alternatively, these associations may highlight shared biological pathways connecting glycaemic regulation with metabolic or inflammatory processes. Future analyses should explore whether these SNPs are vertically pleiotropic by performing via MR mediation analyses.

### Strengths

One of the strengths of this study is that it used UK Biobank, one of the largest sources of imaging samples in the world. Our MR approach generated strong genetic instruments especially for FPG and IR (as per the F-statistics which were 107 and 89 respectively) which gives us confidence in the findings.

Another important strength of our study is that we performed a replication analysis in a different dataset and took a two-sample MR approach using GWAS summary statistics from non-overlapping samples for our glycaemic exposures and brain health outcomes. Using independent datasets gives us more confidence in our effect estimates by eliminating biases like the Winner’s curse and weak instrument bias and increasing our statistical power(Lawlor, 2016). Non-overlapping samples also reduce confounding, as each GWAS sample has independent trait distributions strengthening causal directionality by mitigating risks of reverse causation(Lawlor, 2016). Additionally, our replication approach allowed for efficient, scalable analysis using publicly available data, offering a cost-effective solution for exploring complex causal relationships between metabolic and brain health.

### Limitations

The UK Biobank is subject to a well-documented "healthy volunteer" bias, where participants tend to be healthier, better educated, and more health-conscious than the general population (Fry et al, 2017) This bias may have impacted our MR analyses by underestimating causal effects, particularly for traits or diseases influenced by lifestyle or socioeconomic factors and limiting the generalisability of findings to less healthy populations. Furthermore, the restricted age range of UK Biobank participants, primarily between 40 and 69 years at recruitment may obscure genetic effects that manifest earlier or later in life, particularly for age-related conditions such as dementia restricting insights into life-course trajectories of these traits.

The imaging subsample of the UK Biobanks may represent a more selective group that may differ systematically from the broader cohort, potentially skewing genetic associations derived from imaging data. If these participants are disproportionately healthier or wealthier, the generalisability of MR findings based on this subset could be compromised. Similarly, the potential misclassification of dementia outcomes in the UKB, which relies on hospital records, death registries, and self-reports, poses challenges for MR analyses.

Misclassification may bias effect estimates and hinder the ability to distinguish between dementia subtypes, such as Alzheimer’s disease versus vascular dementia, reducing the reliability of findings (Lyall et al, 2022).

Finally, despite the UKB’s overall large sample size, the limited number of cases for certain outcomes, such as AD subtypes, results in low statistical power for MR analyses of rare or specific conditions. This limitation increases the risk of false negatives and restricts the ability to perform meaningful subgroup analyses. Together, these factors emphasize the importance of interpreting MR findings within the context of these constraints and validating results through external datasets when possible.

We focused on White Europeans since the number of dementia cases in other ancestries was low. Future studies should also investigate the associations we report here in other ethnic groups to ensure that findings are generalisable, accounting for genetic diversity and varying disease risks across populations. Cognitive decline is also an important outcome that we did not investigate, but there would have been very few individuals for this analysis, as only a subsample underwent repeat cognitive testing.

## Conclusions

In this UKB MR study which explored the genetic risk of a range of glycaemic markers with brain health, we found causal relationships between postprandial hyperglycaemia and higher AD risk and fasting insulin and higher overall brain volumes. The former finding was specific to UKB thus future studies are needed to confirm the finding and, if observed, better understand the precise mechanisms through which this may occur. The analyses should also explore whether this association manifests in non-European samples. Future studies should aim to disentangle the mechanisms through which insulin may exert a positive for brain health.

## Data Availability

UK Biobank data was openly available before the initiation of the study and they can be located by the following link: https://biobank.ndph.ox.ac.uk/showcase/search.cgi

## Data Availability Statement

UK Biobank data was openly available before the initiation of the study, and they can be found by the following link: https://biobank.ndph.ox.ac.uk/showcase/search.cgi

